# Development of the Short Hospitalization Predictor (SHoP) Machine Learning Model Across Two Hospitals

**DOI:** 10.1101/2025.05.03.25326935

**Authors:** Richard K. Leuchter, Valiollah Salari, Eilon Gabel

## Abstract

**Objective:** To develop and evaluate an open-source machine learning (ML) models for predicting hospital short stays (length of stay [LOS] under 48 and 72 hours) exclusively using data available at the time of ED admission, with a novel application of target encoding diagnostic codes.

**Materials and Methods:** We trained two ML algorithms (Random Forest and XGBoost) on electronic health record (EHR) data from two hospitals to predict hospital short stays. We employed an innovative weighted target encoding method that converted categorical International Classification of Disease (ICD- 10) codes into numeric representations of their probabilistic contribution to LOS. We measured area under the receiver operating characteristic curve (AUC) for correctly predicting LOS under 48 or 72 hours, which we compared to logistic regression.

**Results:** The final sample included 8,693 adult patients admitted to an internal medicine service. Random Forest models achieved the highest performance for predicting LOS under 48 hours (AUROC=0.96, 95% CI 0.95-0.97; accuracy=91%) and under 72 hours (AUROC=0.94, 95% CI 0.93-0.95; accuracy=88%). These models outperformed logistic regression using the same features (48-hour AUROC=0.57, 95% CI 0.54-0.59 and accuracy=70%; 72-hour AUROC=0.59, 95% CI 0.57-0.61 and accuracy=56%).

**Discussion:** Leveraging an innovative target encoding method, the Short Hospitalization Prediction (SHoP) model substantially outperforms previous ML approaches in accurately predicting LOS under both 48 and 72 hours using only ED pre-admission data (AUC 0.94-0.96).

**Conclusion:** The technical innovation and predictive capability of the SHoP model enables powerful, real-time applications for optimizing patient flow and hospital resource utilization by identifying potentially divertible admissions while patients are still in the ED.

## INTRODUCTION

The U.S. is facing a critical hospital bed shortage as soon as 2032 unless novel care models are developed to reduce the number of hospitalizations.^1^ To mitigate this potential crisis, the management of 10-20% of hospitalizations could be shifted from hospitals to office-based settings,^2–4^ but there are currently no methods to accurately identify these patients being admitted while they are still in the emergency department (ED) triage. A requisite step in designing care pathways that could divert ED admissions to alternative settings is the ability to predict in real-time if a hospitalization will be necessary while a patient is in ED triage.

While directly adjudicating the necessity of hospitalizations is challenging and often subjective, hospital length of stay (LOS) has been proposed as a proxy for divertible hospitalizations.^5^ Specifically, hospital short stays (i.e., LOS under 48 or 72 hours) may suggest that a patient could have been cared for in an alternative setting such as a high-acuity clinic.^5–7^ While machine learning (ML) models have been used to predict LOS with limited success, none have been developed that can accurately predict a LOS under 48 or 72 hours among general medicine patients in ED triage.^8–16^

This study sought to develop open-source predictive models ultimately capable of identifying patients at ED triage who could be diverted to alternative treatment settings. By only training models on data available prior to ED admission, we aimed to create a tool suitable for future real-time implementation, enabling the identification of potentially divertible hospitalizations and subsequent improvements in clinical care and health system operational efficiencies.

## METHODS

### Data Source and Study Population

This retrospective observational study analyzed electronic health record (EHR) data from two academic hospitals (Ronald Reagan UCLA Medical Center and Santa Monica UCLA Medical Center) between March 17, 2021-August 22, 2024. Ronald Regan UCLA Medical Center represents a quaternary care academic hospital caring for medically complex patients including allograft recipients, whereas Santa Monica UCLA Medical Center is more representative of the features of a standard community hospital (less complex case-mix, ED operates without residents, etc.). We identified the records of all patients admitted through the ED at either hospital during the study period. Patients were included if they were at least 18 years old and admitted to an internal medicine service for one of the conditions in eTable 1 of the Supplement, which encompass 16 of the 20 most common reasons for admission in the U.S.^17^ The criteria for primary diagnosis utilized the admitting diagnosis inputted by the ED provider when they placed a bed request for admission, and therefore represented the working diagnosis just prior to hospital admission. Patients were excluded if they were admitted to a surgical, obstetric, neurology, psychiatric, or critical care service.

We utilized only the following de-identified EHR data that were available to ED providers prior to the time of the Admissions Order: basic encounter information, sociodemographics, labs/vitals, and prior healthcare utilization.

The UCLA IRB determined that this secondary analysis of de-identified data did not constitute human subjects research so did not require review.

### Feature Selection

Feature importance scores from the ensemble model were computed in Python using the relevant library (e.g., sklearn for Random Forest or xgboost for XGBoost) to rank features by their contribution to model performance, and low-importance features were removed to optimize the final feature set for training Data missingness after target encoding was completed on the final feature list was less than 2.2%; all missing observations were replaced with -1 which generally indicated a patient had no EHR records at our institution (e.g., -1 as the minimum ED acuity in the last 12 months indicated a new patient with no prior ED visits).

### Target Encoding ICD-10 Codes

Target encoding was chosen as the primary method for processing categorical ICD-10 codes to capture their relationship with LOS in a statistically meaningful way.^19–21^ This approach allowed us to convert categorical values into numeric representations that directly reflected their probabilistic contribution to LOS while mitigating issues such as sparsity and high dimensionality, which are common in healthcare datasets with many categorical variables.^22,23^

To prepare the data for target encoding, a maximum of 25 ICD-10 codes billed during encounters over the prior 12 months per patient were selected, sorted by frequency of occurrence, and organized into separate columns (ICD_CODE_1 to ICD_CODE_25). Approximately 13% of patients had fewer than 25 ICD codes in their charts prior to arrival in the ED so the most common code was repeated in the available spaces until all 25 positions were populated. For the 10% of patients who had no historical ICD codes, the values were filled with historical weighted averages for patients without ICD codes using target encoding.

We then created an encoded value for each ICD-10 code computed as the weighted average of the observed LOS for that diagnostic code and the global average LOS across all observations. We used a weighted averaging scheme to ensure balanced representation of ICD-10 codes, which prevented overfitting to rare categories while still allowing frequent categories to dominate their contribution. The formula used for smoothing encoded values according to weighted averages was:

Encoded Value (weighted) = (Encoded Value X Item Count) + (λ X Global Average)

- Encoded Value: The value representing the average LOS for a given diagnostic code within a particular category of data element (i.e., male v. female sex)
- Item Count: The number of observations for the category.
- λ: A smoothing factor that controlled the contribution of the global average (set at 0.1).
- Global Average: The mean encoded value across all categories of data elements (i.e. the mean LOS for a given diagnostic code for all patients, regardless of sex).

More common ICD-10 codes had much larger weighted encoded values and correspondingly little influence from the global average, whereas less common ICD-10 codes were pulled up further towards the global mean. Target encoded values for each patient’s top 25 ICD-10 codes were used to train the models as 25 independent features.

### Analysis

We used two machine learning algorithms (XGBoost and Random Forest)^24–26^ to categorically predict LOS less than either 48 or 72 hours, compared to logistic regression. Models were trained on a 70% random split of the data and tested on the remaining 30%. Model performance was assessed using the area under the receiver operating characteristic curve (AUC), Precision-Recall AUC (PR-AUC), and accuracy. A Shapley Additive Explanations (SHAP) analysis was conducted to assess feature importance.^27^ 10-fold cross validation was used to validate model performance. All target encoding and ML model training and testing was completed in Scikit-learn Python.^28^

## RESULTS

The final sample included 8,693 patients who were admitted through the ED between March 17, 2021-August 22, 2024. The models were trained on a demographically diverse population of patients who were 35% non-Hispanic White, 24% Hispanic or Latino, 12% Black or African American, and 9% Asian. The remaining sample characteristics are displayed in Table 1, and were balanced between the training and test data sets.

**Table 1.** Sample Characteristics.

|  | Training Set (n=6,092) | Test Set (n=2,601) |
| --- | --- | --- |
| Mean age, years (SD) | 62.0 (21.0) | 62.4 (20.7) |
| No. female (%) | 2,978 (49%) | 1,281 (49%) |
| ***Race and Ethnicity, No. (%)*** |  |  |
| Asian | 539 (9%) | 230 (9%) |
| African American or Black | 721 (12%) | 304 (12%) |
| Hispanic or Latino | 1,455 (24%) | 575 (22%) |
| Non-Hispanic White | 2,159 (35%) | 954 (37%) |
| Unknown or declined | 723 (12%) | 307 (12%) |
| Other | 495 (8%) | 231 (9%) |
| ***Outcomes*** |  |  |
| Mean length of stay, hours (SD) | 135.4 (225.1) | 130.8 (191.6) |
| Length of stay <48h, no. (%) | 1,663 (27%) | 754 (29%) |
| Length of stay <72h, no. (%) | 2,840 (47%) | 1,241 (48%) |

A total of 25 features plus 25 target encoded ICD-10 codes per patient (for a total of 50 features) were used to train the models (Table 2).

**Table 2.** Selected features used to train the Short Hospitalization Prediction (SHoP) Model.

| Feature Name | Description | Feature Importance for Random Forest Model |
| --- | --- | --- |
| ICD_CODES_COMBINED | Target encoded ICD-10 codes from a patient's problem list | 0.7501 |
| IDEAL_BODY_WEIGHT_KG_DELTA | Difference between last recorded weight and calculated ideal body weight based on last recorded height | 0.0227 |
| WEIGHT_KG | Last recorded patient weight | 0.0219 |
| PAT_ZIP | Patient zip code | 0.0215 |
| AGE | Patient age in years | 0.0211 |
| LOS_DAY_IN_LAST_YR | Cumulative hospital bed-days occupied in prior 12 months | 0.021 |
| HOUR_OF_DAY | Hour of the day that the Plan to Admit order was placed | 0.0178 |
| HEIGHT_IN | Last recorded patient height | 0.0156 |
| RACE | Racial category recorded in demographics section of EHR, which is obtained by self-report. Broken down by following categories: African American or Black, Asian, Hispanic or Latino, non-Hispanic White, other, or unknown. | 0.0132 |
| WEEKDAY | Categorical variable indicating the day of the week | 0.0131 |
| PAYOR_NAME | Name of benefit plan, comprised of over 1,400 unique plan names in the training data set. | 0.0125 |
| ED_ADMSN_12_MO | Cumulative number of admissions through the ED in prior 12 months | 0.008 |
| FIN_CLASS | Categorical variable for the following payor types: commercial, group health plan (commercial), international payor, Medicaid, Medicaid-assigned, Medicare, Medicare Advantage, Medicare-assigned, self-pay, Tricare, other). | 0.008 |
| MARITALSTATUS | Marital status recorded in demographic section of EHR, which is obtained by self-report | 0.0073 |
| ADMSN_12_MO | Cumulative number of total admissions (planned and unplanned) in the prior 12 months | 0.006 |
| ADMSN_9_MO | Cumulative number of total admissions (planned and unplanned) in the prior 9 months | 0.006 |
| ED_ADMSN_6_MO | Cumulative number of admissions through the ED in prior 6 months | 0.005 |
| ED_ADMSN_9_MO | Cumulative number of admissions through the ED in prior 9 months | 0.0049 |
| PRIM_LANGUAGE | Primary language recorded in demographic section of EHR, which is obtained by self-report | 0.0048 |
| ADMSN_6_MO | Cumulative number of total admissions (planned and unplanned) in the prior 6 months | 0.0043 |
| ADMSN_3_MO | Cumulative number of total admissions (planned and unplanned) in the prior 3 months | 0.0041 |
| ETHNICITY | Self-reported ethnicity recorded in the EHR. | 0.0038 |
| SEX | Biological sex reported in the EHR. | 0.0033 |
| ED_ADMSN_3_MO | Cumulative number of admissions through the ED in prior 3 months | 0.003 |
| MAX_ED_ACUITY_LAST_12M | The maximum ED acuity score that the patient presented with the ED with in the prior 12 months (scale 1 to 5, 1 is most acute) | 0.0023 |
| MIN_ED_ACUITY_LAST_12M | The minimum ED acuity score that the patient presented with the ED with in the prior 12 months (scale 1 to 5, 1 is most acute) | 0.0021 |

The Random Forest model demonstrated similar performance compared to XGBoost and greatly superior performance compared to logistic regression for predicting hospital short stays under 48 hours and 72 hours (Figure 1). For predicting LOS under 48 hours, Random Forest achieved an AUC of 0.96 (95% CI 0.95-0.97) and an accuracy of 91%, compared to an AUC of 0.96 (95% CI 0.95-0.97) and accuracy of 90% for XGBoost, and an AUC of 0.57 (95% CI 0.54-0.59) and accuracy of 70% for logistic regression. For predicting LOS under 72 hours, Random Forest exhibited an AUC of 0.94 (95% CI 0.93-0.95) and an accuracy of 88%, XGBoost achieved an AUC of 0.94 (95% CI 0.93-0.95) and accuracy of 86%, and logistic regression achieved an AUC of 0.59 (0.57-0.61) and an accuracy of 56%.

**Figure 1.**
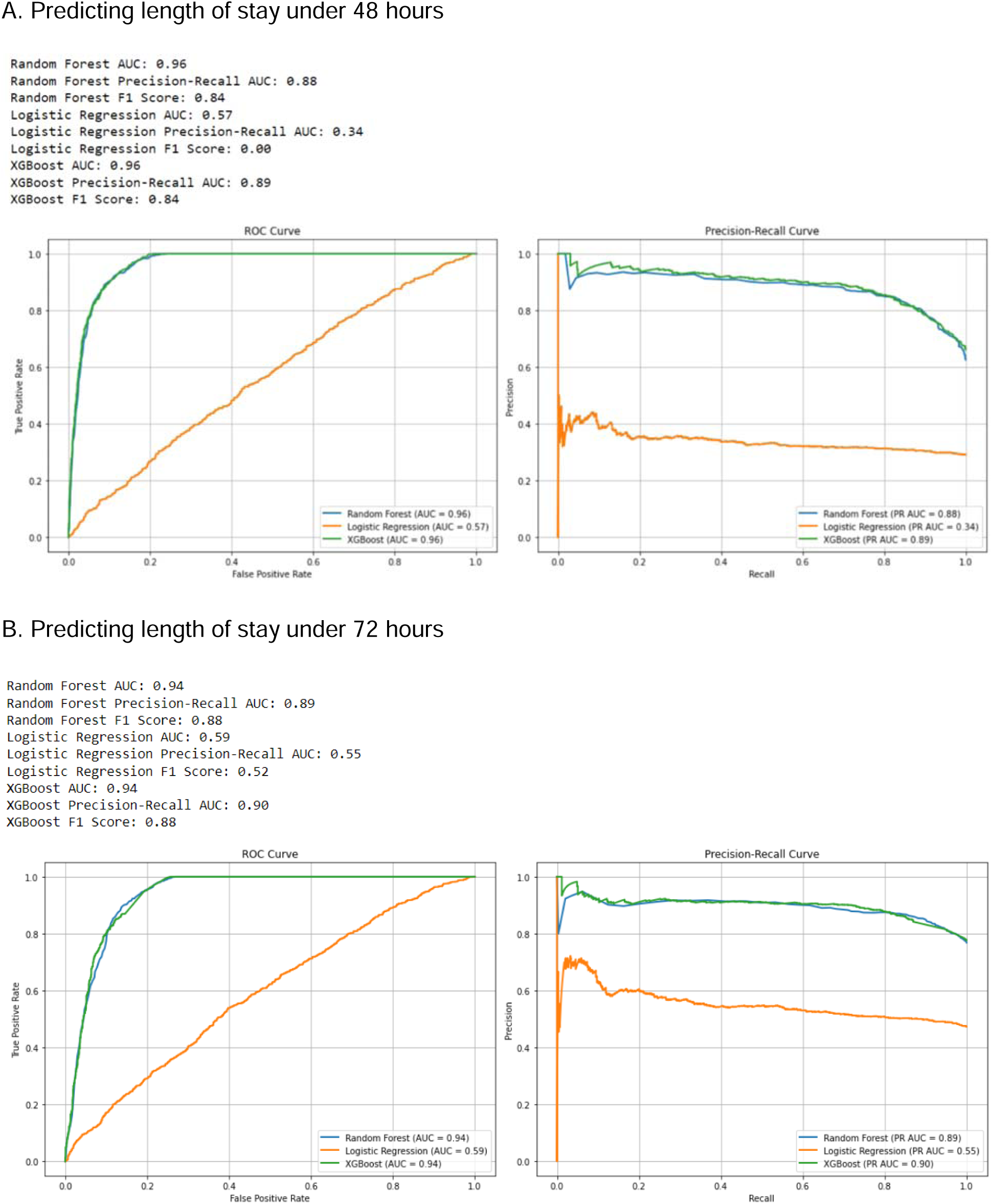
Short Hospitalization Predictor (SHoP) model performance using 48 or 72 hour length of stay thresholds. Figure 1 Legend: The receiver operating characteristic (ROC) curve of each panel depicts the AUC for correctly predicting a length of stay (LOS) under either 48 or 72 hours. The precision-recall curve of each panel depicts a visual representation of the trade-off between the true positive rate (i.e., precision) and positive predictive value (i.e., recall) across a spectrum of thresholds. Each curve compares the indicated metrics between the two machine learning techniques (Random Forest and XGBoost) and logistic regression.

Relative feature importance by SHAP analysis demonstrated that target encoded ICD-10 codes had the largest impact on model output (Figure 2).

**Figure 2.**
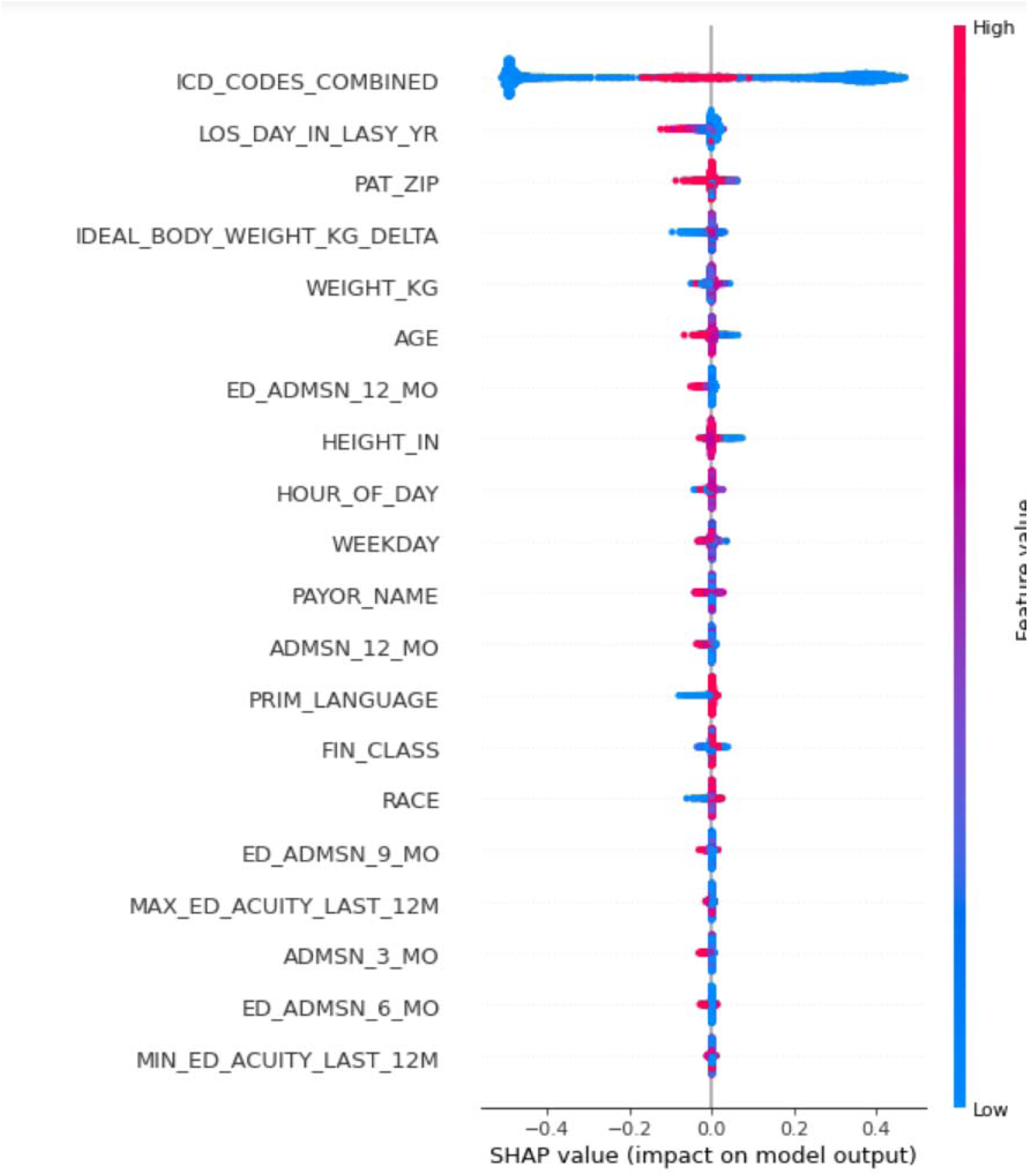
Shapley Additive Explanation (SHAP) value plots for Random Forest Short Hospitalization Predictor model, displaying the top 20 features. Figure 2 Legend: Within each feature row, a single point corresponds to one encounter. Each point’s SHAP value (x-axis) corresponds to the impact that feature had on the model output. This can be interpreted as the indicated feature’s importance for predicting length of stay (LOS). Larger positive SHAP values are associated with longer LOS, whereas larger negative SHAP values are associated with shorter LOS. The color corresponds with the value of the feature itself, with red indicating larger values (e.g., higher age) and blue representing smaller values (e.g., lower age). Features are organized along the y- axis in descending order of explanatory importance.

Model performance was similar across sex and racial/ethnic subgroupings. Accuracy for predicting LOS under 72 hours was as follows: male, 88%; female, 87%; Asian, 91%; Black or African American, 80%; Hispanic or Latino, 87%; non-Hispanic White, 87%; other, 82%.

The code for developing and training the Random Forest and XGBoost models in Python is freely available on Github at the following url: https://github.com/vsalari/NextDayClinic.

## DISCUSSION

This study demonstrates the successful creation of the Short Hospitalization Prediction (SHoP) model using a Random Forest ML algorithm to accurately predict hospital admissions with a LOS under either 48 or 72 hours. The exceptionally high AUC of 0.96 and accuracy of 91% for the 48-hour prediction suggests that the use of target encoded ICD-10 codes is a substantial improvement over prior efforts to use ML models for similar applications.

A key implication of this research is the potential to identify avoidable or divertible hospitalizations in real-time, since the SHoP model exclusively uses a working ED diagnosis and data available before admission. While directly assessing the necessity of each admission is complex, short LOS, particularly those under 48 or 72 hours, can serve as a proxy for potentially divertible admissions that could have been cared for in a high-acuity outpatient setting.^5^

Several studies have explored using ML to categorically predict LOS thresholds, and our findings corroborate the potential of ML for this task.^29^ However, our study makes several important contributions. First, the SHoP model is highly generalizable since it was trained on adult general internal medicine patients being admitted for 16 of the 20 most common admission diagnoses in the U.S., which account for roughly half of all non-obstetric hospitalizations (only osteoarthritis, spondylopathies, depressive disorders, and schizophrenia were not represented due to excluding surgical and psychiatric patients).^17^ A majority of prior models have been trained on specific subpopulations (e.g. cardiovascular or endocrinology patients, older adults)^8–11,13,15^ limiting their utility, especially within an ED setting where diagnoses are often not fully differentiated. Second, the SHoP model notably outperforms previous efforts to predict LOS using ML. Similar models trained on diverse general internal medicine populations have exhibited AUCs ranging from 0.81-0.86,^14,16^ and even models trained on more homogenous groups of patients have attained AUCs from 0.76-0.90 or accuracies between 64-80%.^8,10,11,14,15,30^ Third, the SHoP model addresses critical barriers in successful hospital diversion initiatives by predicting short stays before a patient is admitted. In contrast, previous models’ predictions have required at least 24 hours of hospitalization, used information unavailable at the time of admission (like using final discharge diagnosis rather than the admitting diagnosis), or predicted distant time horizons that do not correlate with avoidable hospitalizations (i.e., under 5, 7, or 14 days).^8,10,11,14,15,30,31^

Importantly, model performance was consistent across sex and racial/ethnic subgroups, with prediction accuracies that did not systematically disadvantage any group despite marginal numerical differences. These findings suggest that, through careful calibration and validation, the SHoP model effectively mitigates bias.

## Limitations

This study has several limitations. First, it is a retrospective study conducted at two hospitals.

Second, model performance is limited in ED admissions with limited or no health system records available at the time of presentation. Third, the SHoP model only considers discrete data stored in the EHR and was not designed to account for data elements such as free text. Fourth, while the SHoP model was trained on a more medically diverse population than most prior models,^8–11,13,15^ it includes only adult general internal medicine patients and excludes cases such as those requiring surgery or critical care.

## Conclusions

To the best of our knowledge, the SHoP model is the first completely open-source model to accurately predict short hospital stays (under 48 and 72 hours) using only data available at the time of ED admission. Compared to prior ML models, the use of target encoded ICD-10 codes in the SHoP model allows it to accomplish a more difficult prediction task (i.e., predicting narrower windows for LOS using limited information) with substantially higher accuracy.

For ease of visualization, all target encoded ICD-10 codes (top 25 most frequently occurring per patient) were compressed into a single feature row.

## Author Contributions

Concept and design: Leuchter, Gabel

Acquisition, analysis, or interpretation of data: Leuchter, Gabel, Salari Drafting of the manuscript: Leuchter

Critical revision of the manuscript for important intellectual content: Leuchter, Gabel, Salari Statistical analysis: Salari and Gabel

Obtained funding: N/A.

Administrative, technical, or material support: Leuchter, Gabel Supervision: Leuchter, Gabel, Salari

## Code Availability

The code is freely available on Github as noted above.

## Data Availability

Data was never abstracted for this study; training and testing of models was done on a secure cloud-based sever that prohibits the authors from extracting data for distribution. Thus, it is not technically feasible to share the training or test data.
The code used to train the models is freely available on Github and open source, as noted in the manuscript text.

https://github.com/vsalari/NextDayClinic

