## Supplement for "Development of the Short Hospitalization Predictor (SHoP) Machine Learning Model Across Two Hospitals"

**eTable 1. List of diagnoses used to train models.**

| **Illness** | **Value Set Authority Center**^42^ **Diagnostic Code Set Source** | **ICD-10 Codes** |
| --- | --- | --- |
| Abdominal pain | CSTE | R14, R10 |
| Acute kidney injury (AKI) | NCQA (value set for kidney failure) | N17, S37.00, N19 |
| Anemia | The Joint Commission | D46.0, D46.2, D46.4, D46.9, D50, D51, D46.1, D52, D53, D55, D59.0, D59.1, D59.2, D59.4, D63, D59.8, D61, D64, D59.9, D59.5, D59.6 |
| Arrhythmia | AHA | I48, I49.2, I49.3, I49.4, I49.5, I49.8, I49.1, I49.9, I47.0, I47.1, I47.9, R00 |
| Asthma | ACEP and AMA | J45, J82.83 |
| Bronchitis | None | J40, J98.01, J98.09, J68.0, J22, J20 |
| Cellulitis | NCQA and CSTE | L02, L98.0, L98.1, L98.3, L98.411, L98.418, L03, L98.419, L98.421, L98.428, L98.429, L98.491, L98.498, L98.499, L98.9, L98.5, L98.8, H60.1, L01, L98.6, K12.2, K61, N73.0, N73.1, N73.2 |
| Chest pain | ACEP and AMA | R07.1, R07.8, R07.9, I20.1, I20.8, I20.9, R07.2 |
| Chronic kidney disease (CKD) | Health Level 7 Patient Care Working Group | N18.1, N18.3, N18.4, N18.9, N18.2, Q61.2, Q61.3, Q61.8 |
| Chronic obstructive pulmonary disease (COPD) | Lantana | J41, J42, J43, J44.0, J44.1, J44.9, J47.1, J47.9, J98.2, J98.3, R06.2 |
| Colitis | None | K52, K51.011, K51.018, K51.019, K51.20, K51.211, K51.00, K51.218, K51.219, K51.31, K51.318, K51.319, K51.40, K51.411, K51.511, K51.418, K51.50, K51.818, K51.518, K51.419, K51.519, K51.80, K51.811, K51.819, K51.90, K51.911, K51.918, K51.919, A04, K50.00, K50.011, K50.018, K50.018, K50.10, K50.111, K50.118, K50.119, K50.80, K50.811, K50.818, K50.819, K50.90, K50.911, K50.918, K50.919, A09, A04.7 |
| Congestive heart failure (CHF) | ACEP and AMA | I11.0, I13.2, I50, E87.70, E87.79, I25.5, I13.0, I42.9, I42.0, I09.81 |
| Coronary artery disease, non-acute coronary syndrome (CAD) | AHA | I25.1 |
| Dehydration | Oncology Nursing Society | E86 |
| Diarrhea | CSTE | R19.7, K58.0, K59.1, A07.9, K59.1 |
| Diverticulitis | None | K57.1, K57.5, K57.9, K57.3 |
| Dyspnea | Change Healthcare | R06.0, R06.3, R06.4, R06.5 ,R06.8, R06.9 |
| Electrolyte abnormality | None | E87.0, E87.2, E87.3, E87.4, E83.41, E83.42, E87.1, E83.51, E87.5, E87.6, E87.8, E83.52 |
| End-stage renal disease (ESRD) | Health Level 7 Patient Care Working Group and Mathematica | N18.6, Z99.2, Z49.0, Z49.3, N18.5, I12.0, I13.11 |
| Fever | CSTE | R50.2, R50.9, A68, R50.8 |
| Gastrointestinal bleed | CTSE (value set for hematemesis) | K21.01, K28.4, K28.7, K29.01, K29.21, K29.31, K22.11, K29.41, K29.51, K29.61, K29.71, K29.81, K29.91, K62.5, K92.0, K92.1, K92.2 |
| Hematuria | Health Level 7 Patient Care Working Group | R31, N30.41, R82.3, N02, N30.11, N30.21, N30.31, N30.01, N30.91 |
| Hypertension | NCQA | I10, I12.9, I16.0, I16.1, I16.9, I13.10, I11.9, I15 |
| Hypotension | AHA | I95, R03.1 |
| Leukocytosis | None | D72.10, D72.82 |
| Lower urinary tract infection (UTI) | NCQA | N28.84, N28.86, N30.00, N30.10, N30.20, N30.30, N30.40, N30.80, N30.90, N34.1, N34.2, N34.3, R30.0, N39.0, B37.4, R82.81 |
| Osteomyelitis | CSTE | M86, M46.3, M46.4, M46.5, A54.43, A02.24, M46.2, H70, H95, M90.8 |
| Pancreatitis | CSTE | K85.00, K85.10, K85.11, K85.20, K85.21, K85.30, K85.01, K85.31, K85.80, K85.81, K85.90, K85.91, K86.0, K86.1, K86.2, K86.3 |
| Pleural effusion | None | J90, J94.0, J91 |
| Pneumonia | Lantana | J12, J14, J15, J16, J18, B37.1, J13, A43.0, A54.84, B59, A37.11, A37.81, A70, A37.01, B39.0, B39.1, B39.2, J68.0, B06.81, A37.91, J18.0, J18.08, J18.9, J69, J82.81, J82.82, J84.111, J84.113, J84.114, J84.116, J84.117, J84.9 |
| Pulmonary fibrosis | None | J84.10, J84.178, J84.9, J84.112 |
| Pulmonary hypertension (PH) | The Joint Commission | I27 |
| Pyelonephritis | None | N10, N12, N16, N15.1, N15.9 |
| Sepsis | ACEP and AMA | R65.2, A26.7, A32.7, A42.7, A41, A22.7, R65.11, T81.44, A02.1, B37.7, T88.0, A48.3, R78.81, A40, A54.86, A02.1 |
| Syncope | None | R55, G90.01, T67.1 |
| Urinary retention | LUGPA | R33, N40 |
| Vomiting and nausea | ACEP and AMA (value set for vomiting), CSTE (value set for nausea) | R11 |
